# The use of social media platforms by migrant and ethnic minority populations during the COVID-19 pandemic: a systematic review

**DOI:** 10.1101/2022.02.07.22270579

**Authors:** Lucy P Goldsmith, May Rowland-Pomp, Kristin Hanson, Anna Deal, Alison F Crawshaw, Sally E. Hayward, Felicity Knights, Jessica Carter, Ayesha Ahmad, Mohammad Razai, Tushna Vandrevala, Sally Hargreaves

## Abstract

**Objective:** To determine the extent and nature of social media use in migrant and ethnic minority communities for COVID-19 information, and implications for preventative health measures including vaccination intent and uptake.

**Design:** A systematic review of published and grey literature following the Preferred Reporting Items for Systematic Review and Meta-Analyses (PRISMA) guidelines

**Eligibility Criteria for study selection:** Global research reporting the use of social media by migrants and/or ethnic minority groups in relation to COVID-19.

**Data extraction:** We extracted data on key outcomes, study design, country, population under study, and sample size.

**Results:** 1849 unique records were screened, and 21 data sources included in our analysis involving migrant and ethnic minority populations in the UK, US, China, Jordan, Qatar, and Turkey. We found evidence of consistent use of a range of social media platforms for COVID-19 information in some migrant and ethnic minority populations (including WeChat, Facebook, WhatsApp, Instagram, Twitter, YouTube), which may stem from difficulty in accessing COVID-19 information in their native languages or from trusted sources. There were positive and negative associations with social media use reported, with some evidence suggesting circulating misinformation and social media use may be associated with lower participation in preventative health measures, including vaccine intent and uptake, findings of which are likely relevant to multiple population groups.

**Conclusions:** Social media platforms are an important source of information about COVID-19 for some migrant and ethnic minority populations. Urgent actions and further research are now needed to better understand the use of social media platforms for accessing health information by different population groups – particularly groups who are marginalised from health systems – effective approaches to tackling circulating misinformation, and to seize on opportunities to make better use of social media platforms to support public health communication and improve vaccine uptake.

**Registration:** This study has been registered with PROSPERO (CRD42021259190).

## Introduction

The pandemic has been accompanied by an infodemic, defined as an excess of information during a disease outbreak – including false or misleading information in digital and physical environments^1^ – that makes it difficult to distinguish reliable information from misinformation including disinformation (deliberate misinformation) and conspiracy theories. The World Health Organization (WHO) highlights that in all communities, infodemics cause ‘confusion and risk-taking behaviours that can harm health…it leads to mistrust in health authorities and undermines the public health response, and can intensify or lengthen outbreaks’^1^. The rapid expansion of internet and social media use, in particular, in recent years (including platforms such as Twitter, WhatsApp, and YouTube; Table 1) has meant that both useful and potentially harmful health information can spread rapidly. A large proportion of the most popular COVID-19 videos on YouTube, for example, have been found to contain misinformation, or no factual information, reaching millions of people worldwide^2 3^. YouTube is considered a major platform for information concerning the control of COVID-19, but most COVID-19 videos were of ‘undesirable quality’ containing few government/public health recommendations according to a recent study^4^. A review of YouTube videos on general vaccination found 65% expressed anti-vaccination sentiment^5^, with anti-vaccine posts more likely to be recirculated on Twitter^2^. The spread of misinformation and disinformation has been highlighted as a major risk to ending the COVID-19 pandemic – including undermining trust in vaccines^6^ – with researchers highlighting links between misinformation on social media and public doubts around vaccine safety, self-reported compliance with public health guidelines, and intent to vaccinate^7^,^8^.

**Table 1:** Popular social media platforms Statistics from Statistica (2021) ^68^.

| Platform | Primary Feature | Country of origin | Organisation | Users |
| --- | --- | --- | --- | --- |
| YouTube | Online video sharing and social media platform. Free to use. | US | Google | Approximately >2 billion monthly |
| WhatsApp | Messaging platform, allows users to send text messages and voice messages, make voice and video calls, and share images, documents, user locations, and other content. Free to use. | US | Meta | Approximately >2 billion monthly |
| Instagram | Photo and video sharing social networking site. Free to use. | US | Meta | Approximately 1 billion monthly |
| Facebook | Social networking service, allows messaging, image and video sharing, marketplace online shopping, live video sharing. Free to use. | US | Meta | 2.89 billion active monthly |
| WeChat | Instant messaging, social media, mobile payment. Free to use. | China | Tencent Holdings Limited | 1.25 billion monthly |
| TikTok<br>(Known in China as Douyin) | Video sharing focused on short form videos (15 seconds – 3 minutes). Free to use. | China | ByteDance | 837 million monthly active |
| Snapchat | Photo sharing multimedia app with video features. Free to use. | US | Snap Inc. | 347.3 million monthly active |
| Twitter | Microblogging focused on short messages known as 'tweets'. Live chat event function Tweetchat. | US | Twitter Inc. | 330 million monthly active |

Although social media platforms are commonly used in the general population, and patterns of use are complex across different population groups^8 9^, some migrant and ethnic minority groups – who may experience barriers to accessing health information and health systems – may be more reliant on social media and the internet as a source of health information. These communities may also draw on diaspora media as a source of health information^10^. The COVID-19 pandemic has disproportionately impacted and exacerbated inequalities faced by migrants and ethnically diverse communities^11 12 10 13 14^, with lower take-up of preventative health measures, such as vaccines, noted in some groups due to a range of personal, societal, and physical barriers^12 14 15^. Some migrant and ethnic minority communities may be more exposed to social media misinformation because of access barriers to accurate information (eg, from official government sources)^16 17^, due to restricted eligibility and access to services, language barriers, and low health literacy. However, little is known about the extent and nature of social media use in these populations, nor the impact that social media use has had on preventative health measures during the pandemic, including COVID-19 vaccine uptake. In addition, there is an opportunity now to explore the extent to which social media platforms could be better used to support information sharing and promote public health messaging in marginalised communities during the pandemic and beyond.

We therefore did a systematic review to explore and assess the extent and nature of social media use by migrant and ethnic minority groups to access COVID-19 health information, the extent to which misinformation on social media may have influenced views about COVID-19 preventative measures including vaccination intention and uptake, and to explore good practice.

## Methods

### Search Strategy

The review was registered with PROSPERO (CRD42021259190)^18^ and followed PRISMA guidelines^19^. A Boolean search strategy was developed containing terms relating to migrants, ethnic minorities, COVID-19, social media, and misinformation (see supplementary information 1). We searched the following databases: Embase, Web of Science, Oxford Academic Journals, PubMed NIH, Clinical Trials, China CDC MMWR, CDC reports, ProQuest Central (Proquest), CINAHL, Africa Wide Information (Ebsco), Scopus, PsycInfo, CAB Abstracts, Global Health, J Stage, Science Direct, Wiley Online Journals, JAMA Network, British Medical Journal, Mary Ann Liebert, New England Journal of Medicine, Sage Publications, Taylor and Francis Online, Springer Link, Biomed Central, MDPI, ASM, PLOS, The Lancet, Cell Press, and pre-print sites chemRxiv, SSRNbioRxiv, and medRxiv facilitated through the WHO Global Research on COVID-19 database from inception to 9/6/2021 (https://search.bvsalud.org/global-literature-on-novel-coronavirus-2019-ncov/). The WHO’s COVID-19 Database^20^, is a daily updated multilingual resource of all literature (peer-reviewed literature, pre-prints and grey literature) pertaining to COVID-19.

Records were imported to Rayyan QCRI^21^. Both title and abstract screening and full text screening were conducted independently by two reviewers (MR-P and LG) using Rayyan QCRI^21^, with discrepancies resolved by a third reviewer (SH). Additional relevant papers and grey literature (e.g. from third-sector organisation websites) were identified using hand searching including backwards and forwards citation tracking.

### Selection criteria and primary outcomes

Papers reporting the use of social media platforms and implications for preventative health measures and vaccination intent of migrants and/or ethnic minority groups to COVID-19 globally were eligible. All types of scientific articles, reports and commentaries, editorials, correspondence letters were eligible for inclusion. Social media platforms were defined as any medium whereby content (including images, videos, and messages) is circulated to the general public and may include YouTube, Facebook, Twitter, TikTok, and Snapchat. ‘Migrants’ were defined as foreign-born, residing outside of their country of birth. An ethnic minority group was defined as a group of people with a shared culture, tradition, language, history, living in a country where most people are from a different ethnic group, and will include migrants/foreign-born populations alongside individuals born in the host country. Where studies reported a general population sample, results about migrant/ethnic minority groups within that sample were eligible for inclusion. No papers were excluded based on language or geographical origin. Studies were excluded if it was not possible to determine whether individual(s) in the population studied were migrants or from an ethnic minority group.

### Data Extraction, critical appraisal, and synthesis

Data extraction was completed independently by two researchers (MRP and LG) using a piloted, structured data extraction sheet in Microsoft Excel and data were collated and assessed using narrative synthesis. Risk of bias was assessed independently by two researchers (LG, MRP) using the Quality assessment for Survey Studies in Psychology for Surveys (Q-SSP)^22^ for quantitative studies. The twenty items on this scale can be rated as “yes”, or “no”, “not stated clearly”, or “not applicable”. Scores are calculated by dividing the “yes” answers by the total number of applicable items, with scores over 70% indicating “acceptable” quality. The Critical Appraisal Skills programme (CASP) checklist was used for qualitative studies^23^. The ten items can be rated ‘yes’, ‘can’t tell’ or ‘no’. We rated the CASP by dividing the “yes” answers by the total number of applicable items, with a score of over 60% indicating “acceptable” quality. We did not exclude any papers on the basis of quality.

## Results

### Overview of data sources

Following de-duplication, 1849 unique data sources were identified and screened and ultimately 129 were full-text screened. 21 data sources were included in the final analysis (Figure 1). Six studies were conducted in the UK^24-29^, two were jointly conducted in the UK and US^30 31^. An additional eight studies were conducted in the US,^32-39^ and one each in China,^40^ Jordan,^41^ Qatar,^42^ and Turkey.^43^ Eight studies reported on migrants,^27 29 32 40-44^ including migrants in the host countries of China^40^, Jordan^41^, Quatar,^42^ Turkey^43^, and the US^32^ and UK^27 29^, and one study involved predominantly migrants from Venezuela residing in other countries.^44^ Nine studies reported about a specific ethnic minority or group (Latino individuals,^32 34 36 37^ Black American citizens,^35 39^, Jain community members^28^ and Syrian migrants^41 43^). Seven studies reported about ethnic minority groups generally^24-26 30 31 33 38^. A survey design was the most common design, used in half of included studies.

**Figure 1:**
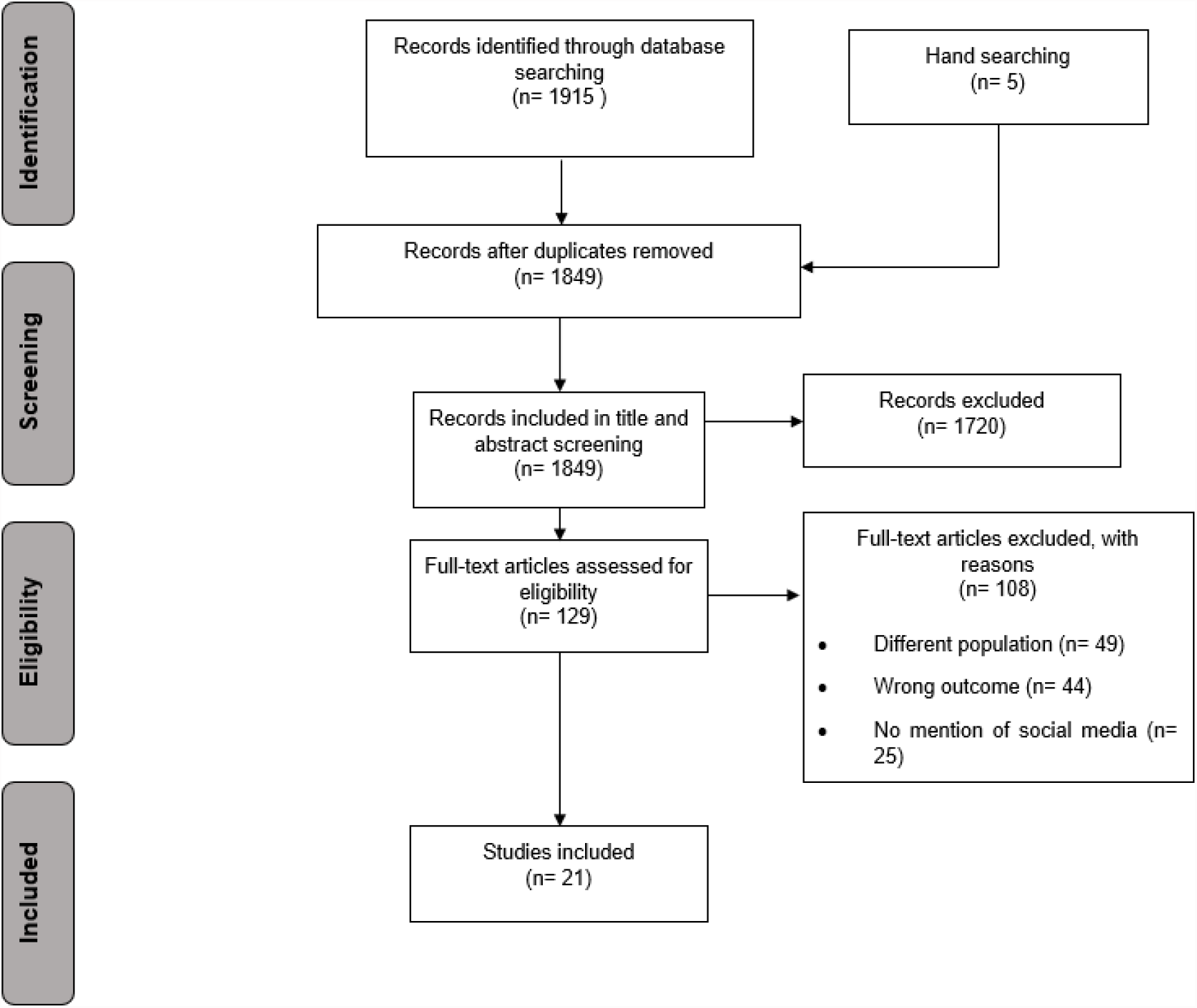
PRISMA diagram of included studies.

Characteristics of included studies are presented in Table 2, including the risk of bias assessment scores. Quality scores ranged from 76% to 90% for included papers, suggesting acceptable quality of all included data sources where quality assessment was applicable. Figure 2 shows the geographical location of data sources, highlighting the absence of published and unpublished data on this topic from most regions of the world.

**Table 2:** Characteristics of included papers.

| **Included study** | **Location of study** | **Study design** | **Population under study** | **Sample size** | **Quality rating\*** |
| --- | --- | --- | --- | --- | --- |
| **Alabdulla 2021<sup>42</sup>** | Qatar | Survey | Non-Qatari residents | 7821 | 76%<sup>1</sup> |
| **Allington 2021(a)<sup>26</sup>** | UK | Survey | Non-White ethnic groups | 4343 | 82%<sup>1</sup> |
| **Allington 2021(b)<sup>31</sup>** | UK | Survey | Non-White ethnic groups | 8988 | 89%<sup>1</sup> |
| **Behbahani 2020<sup>32</sup>** | US | Organisational case study | Latino migrants | N/K | N/A<sup>1</sup> |
| **Campos-Castillo 2020<sup>33</sup>** | US | Survey | Non-White ethnic groups | 10,510 | 88%<sup>1</sup> |
| **Cervantes 2021<sup>34</sup>** | US | Qualitative interviews with thematic analysis | Low-income Latino individuals | 60 | 90%<sup>3</sup> |
| **Chandler 2020<sup>35</sup>** | US | Qualitative interviews with thematic analysis | Black women (18-31yrs) | 15 | 90%<sup>3</sup> |
| **Crawshaw 2021<sup>27</sup>** | UK | Evidence synthesis linked to outputs from participatory workshops with migrants | International migrants | N/K | N/A<sup>2</sup> |
| **Despres 2020<sup>36</sup>** | US | Organisational case study | Latino community living in America | N/K | N/A<sup>2</sup> |
| **Danish Refugee Council (DRC) 2020<sup>43</sup>** | Turkey | Survey | Syrian refugees in South-East Turkey | 774 | 82%<sup>1</sup> |
| **Hamadneh 2021<sup>41</sup>** | Jordan | Survey | Syrian refugee mothers | 389 | 78%<sup>1</sup> |
| **Lockyer 2021<sup>24</sup>** | UK | Qualitative interview; reflective thematic analysis | People from different ethnic groups in Bradford | 20 | 90%<sup>3</sup> |
| **Loomba 2021<sup>30</sup>** | UK & USA | Randomised controlled experiment | Other ethnic groups than White | 4,000 (UK)<br>4,001 (US) | N/A<sup>2</sup> |
| **Moyce 2020<sup>37</sup>** | US | Qual interviews narrative synthesis | Latino individuals | 14 | 90%<sup>3</sup> |
| **Paul 2021<sup>25</sup>** | UK | Repeated measures survey; cohort study | Other ethnic groups than White | 32,361 | 89%<sup>1</sup> |
| **Regional Inter-agency coordination platform (R4V) 2021<sup>44</sup>** | Any host country for migrants from Venezuela | Survey | Predominantly migrants from Venezuela | 334 | 90%<sup>3</sup> |
| **Vekemans 2021<sup>28</sup>** | UK | Organisational case study | Jain community members | 25,000 estimate | N/A<sup>2</sup> |
| **Viswanath 2021<sup>38</sup>** | US | Survey | Non-White ethnic groups | 1012 | 78%<sup>1</sup> |
| **Wang 2020<sup>40</sup>** | China | Survey | International migrants | 1,426 | 78%<sup>1</sup> |
| **Woko 2021<sup>39</sup>** | US | Survey | Black American citizens | 1,074 | 77%<sup>1</sup> |
| **Deal 2021<sup>29</sup>** | UK | Qualitative in-depth interview study | Precarious migrants (asylum seekers, undocumented migrants, refugees) | 32 | 90%<sup>3</sup> |

\*Scores were calculated on both scales by dividing the “yes” answers by the total number of applicable items.
<sup>1</sup> Quality assessment for Survey Studies in Psychology for Surveys (Q-SSP) Checklist for surveys
<sup>2</sup> N/A = not applicable due to research item design.
<sup>3</sup> Critical Appraisal Skills programme (CASP) checklist N/K=not known.

**Figure 2:**
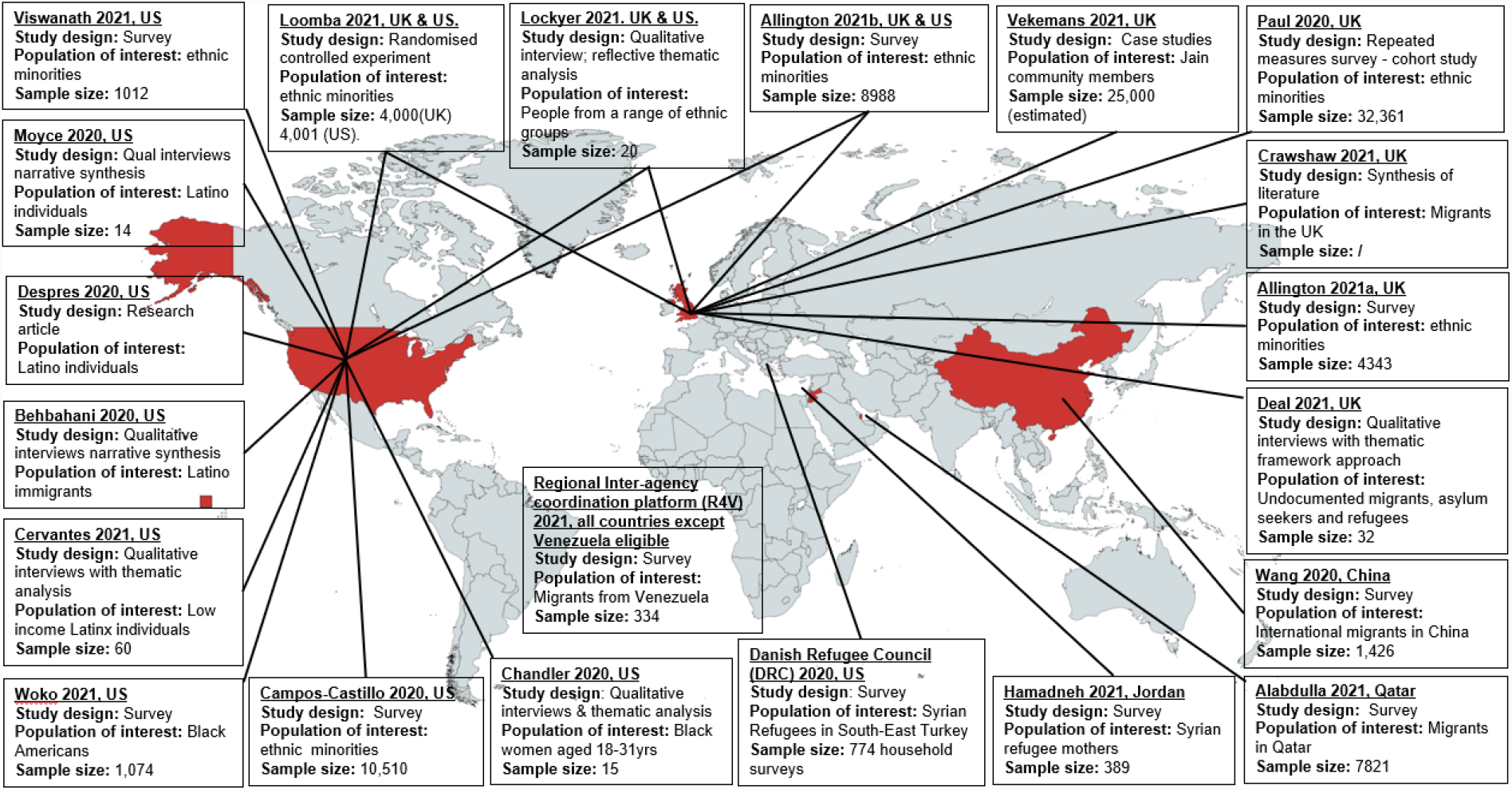
Regional distribution of included data sources.

### Use of social media platforms as a source of information about COVID-19

For some migrants and ethnic minority groups, consistent use of social media platforms for sharing and receiving COVID-19-related health information was reported in several included studies^29 34-36 40-42 44 43^. Social media was reported to be the preferred source of information about COVID-19 for international migrants in China (WeChat was used by 94.5% of respondents for COVID-19 information).^40^ Among 389 Syrian refugee mothers in Jordan^41^, Facebook and WhatsApp were the main sources of information for 87% and 69% of respondents respectively for COVID-19 information; with 21% indicating that they accessed information from professional databases or government websites, and 53% via television (this survey was circulated via Facebook and WhatsApp). Migrants from Venezuela (residing in numerous countries) reported Facebook and WhatsApp were their two primary sources of information about COVID-19 in a survey of 334 migrants^44^. A survey of 774 refugee households in Southeast Turkey^43^ found the majority (75%) obtained COVID-19 information from Facebook, YouTube, Twitter and the internet in general, 15% via SMS/WhatsApp messages, followed by radio/TV (64%) and members of their community/family (34%); only 10% reported getting information from NGO/UN sources. This study concluded that the heavy reliance on social media for information may expose a sizeable proportion of refugee households to fake or inaccurate information. In a US study of Black women aged 18-31 years, 58% of respondents reported using social media (Instagram and Facebook) to obtain COVID-19 information^35^. Participants from the US Latino community described relying on social media for information about the pandemic^34^. In Qatar, migrants reported they preferred to find out about COVID-19 using their own personal research or searching for information, including using social media as a source^42^. A study of precarious migrants (asylum seekers, undocumented migrants) in the UK found many relied on social media (WhatsApp groups, Facebook) for information on the pandemic and the ongoing vaccination programme^29^.

A key theme emerging in one UK study of ethnic minority groups^24^ was that the “avalanche” of information surrounding COVID-19 had led to interviewees feeling overwhelmed and confused: participants reported using a variety of sources of information, including TV, radio, news stations in Pakistan, India, Slovakia, and Poland, online newspapers, Facebook, WhatsApp, Twitter, Google, and medical journals. A number of these participants said they dismissed some stories encountered on WhatsApp and Facebook; however, the sheer volume of messages coupled with the fact that people they trusted were sharing them, proved difficult to ignore, with participants raising concerns about how quickly social media stories were shared. One study exploring the views of US Latinos reported that they consulted national and local news reports for information about COVID-19 and many reported that they got their news from Spanish-language news due to difficulty in understanding news in English; some received their news from social media sources, including Facebook, but expressed caution around messages from social media as there was no way to ensure the accuracy of the reports.^37^

According to one study, member of ethnic minority groups were also more likely to post COVID-19 content on social media than White individuals^33^, with respondents who identified as Black (odds ratio [OR] 1.29, 95% CI 1.02-1.64; P=.03), Latino (OR 1.66, 95% CI 1.36-2.04; P<.001), or other races/ethnicities (OR 1.33, 95% CI 1.02-1.72; P=.03) had higher odds than respondents who identified as White of reporting posting COVID-19 content on social media.

### Drivers of social media reliance

Studies reported that some migrant and ethnic minority groups turned to social media due as a result of a need for connection and to acquire accessible information from people they considered to be reliable sources. For the Latino community in the US, faith and community bonds were valued ways of coping with the difficulties of the pandemic which included feelings of social isolation, stress, and uncertainty and – according to one study – social media facilitated these connections in a virtual space^37^. The Jain community in London used social media to communicate news and knowledge about COVID-19 and stay connected online, with events moving to a virtual space; individuals reportedly benefited from and were grateful for this community use of social media^28^.

Several studies highlight concerns that some migrant and ethnic minority groups were unable to find official information in their host country in their native language about various aspects of COVID-19, hence their reliance on social media^24 27 29 40 43^. For example, a UK study of precarious migrants (asylum seekers, undocumented migrants) reported that those feeling most abandoned or scared due to a lack of understandable, clear official information in the early stages of the pandemic were more likely to rely on word-of-mouth or social media (WhatsApp groups, Facebook) for information, including around the vaccination programme^29^. One study of international migrants in China (94.5% of whom preferred social media for news about COVID-19) had lower rates of correct knowledge about COVID-19 compared to rates reported for Chinese residents^40^. The authors speculate that this might be due to a lack of available public health information in a range of languages.

Other studies showed positive associations with use of social media and access to information. One study highlighted that social media can support migrants to navigate the complex medicolegal context of their host countries by accessing information about public health measures and how to access medical help^32^. Social media use was associated with improved knowledge about COVID-19 and how to stay safe, in studies of Syrian refugee mothers^41^ and US Latinos^36^. In another study specifically curated, culturally relevant digital content was considered to be an effective health promotion tool to share knowledge about practical actions to be taken to address the inequitable impact of the pandemic on US Latinos^36^.

### Misinformation and social media use

A summary of some of the key misinformation narratives identified in studies are provided in Table 3. Some studies made links between social media and circulating misinformation in migrant and ethnic minority groups. For example, a UK cohort study found that both belonging to an ethnic minority group and socioeconomic disadvantage was associated with both exposure to misinformation about vaccines, and mistrust in information about COVID-19^25^. A study of Syrian refugee mothers in Jordan, who reported receiving most of their COVID-19 information through social media, identified some erroneous beliefs about pregnancy, COVID-19 and breast milk^41^. A UK study among ethnic minority groups reported that participants encountered a range of misinformation, usually through social media sources and that vaccine hesitancy could be attributed to safety concerns, negative stories and personal knowledge, all of which had been amplified by recent exposure to misinformation via social media^24^. Myths identified included the idea that health professionals at the local hospital were injecting people with COVID-19 or killing people with the COVID-19 vaccine; there were wider beliefs reported about vaccines containing a microchip; making people infertile, or that vaccines are being tested on ethnic minority individuals^24^. These participants described the dilemma of not knowing what to trust or who to listen to, including the videos /posts that appeared to be from trusted professionals; therefore, they could not entirely dismiss negative stories circulating via social media and elsewhere.

**Table 3:** Examples of circulating misinformation on social media platforms relating to COVID-19 (2020), reproduced and compiled from Loomba et al^30^ and Lockyer et al.^24^.

| Misinformation identified | Source | Engagement<sup>1</sup> | Reach<sup>2</sup> |
| --- | --- | --- | --- |
| "Scientists have expressed doubts over the effectiveness of a COVID-19 vaccine that has been rushed to human trials, after all the monkeys used in initial testing later contracted COVID-19." | Twitter | 1.59K | 1.5m |
| "The new vaccine for COVID-19 will be the first of its kind EVER. It will be an mRNA vaccine which will literally alter your DNA. It will wrap itself into your system. You will essentially become a genetically modified human being" | Twitter | 27 | 19.6K |
| "They said it was just to flatten the curve. Now it's a battle for human survival." The only must-see action thriller for 2020. Starring: Bill Gates, Anthony Fauci, Chris Witty, Matt Hancock. Guest mask appearances: Clintons, Boris Johnson, Nicola Sturgeon, Joe Biden & Tedros. [Graphic featuring Mr. Bill Gates with the following quote.] "If we do a really good job | Twitter | 11 | 1.49K |
| with vaccines, we can reduce population by up to 15%. But if we create a worldwide pandemic first, killing people and making many of the survivors sterile, then create the vaccine, we may achieve the Georgia Guidestones 1st commandment!" |  |  |  |
| Something is very fishy about all this indeed. "A VIRUS WITH A 99.6% SURVIVAL RATE FOR PEOPLE UNDER 70 BUT THE ENTIRE WORLD NEEDS TO TAKE A VACCINE? I'M NO SHERLOCK HOLMES BUT SOMETHING'S FISHY ABOUT ALL THAT....." | Twitter | N/K<sup>2</sup> | 32.5K |
| "Big Pharma whistle-blower: '97% of corona vaccine recipients will become infertile'" | Twitter | 6.95K | 336K |
| "I've been in Twitter jail for the last 12 hours for posting a link to a peer reviewed scientific study published in Vaccine showing that in military personnel prior receipt of the flu shot increased COVID-19 risk by 36%. Censorship is vile & unAmerican." | Twitter | 25.1K | 1.41K |
| "So we know for a fact that the flu vaccine worsens COVID-19 symptoms. So what are they mandating now? The flu vaccine, of course." | Facebook | NK | NK |
| "PREPARING THE PROPAGANDA BLITZ. Yale University and the U.S. government are running clinical trials to develop propaganda messaging to persuade Americans to take experimental, genetically engineered, unlicensed, "Warp Speed," zero liability, expedited vaccines with limited short duration safety testing. Researchers compared reactions in 12 focus groups using "guilt, embarrassment, bravery, anger, trust" and "fear" to overcome vaccines hesitancy" | Instagram | 28.2K | NK |
| COVID-19 is not real, it is an effort to control society<br>COVID-19 has been manufactured by China or other governments for control purposes | Multiple platforms/unknown | NK | NK |
| COVID-19 is caused by 5G<br>COVID-19 has been invented to make people use contactless payments so that the government can track individuals<br>COVID-19 testing gives so many false positives that it is ineffective and you should not self-isolate<br>COVID-19 exists but is not as virulent as the government says it is<br>If children test positive for COVID-19 during school hours, they can be taken away into care and will not be able to see their parents until they test negative<br>The COVID-19 vaccine contains a chip that will track individuals, stop them travelling etc<br>The COVID-19 vaccine will make people infertile and is an attempt to reduce the population, particularly targeted at people from BAME communities<br>BAME people are being used as 'guinea pigs' to test out the COVID-19 vaccine<br>The COVID-19 vaccine has been developed and approved too quickly and has not been fully tested<br>The COVID-19 vaccine will negatively disrupt your natural immune system<br>Herbal remedies will be more effective than the COVID-19 vaccine |  |  |  |

1 Engagement measures the number of likes and retweets.
2 Reach measures the number of followers and thus potential audience size.
NK=Not known.

In a study of 60 Latinx adults hospitalised for COVID-19 in the US, many participants reported that they relied on social media for COVID-19 recommendations and described a lack of information and circulating misinformation, with suspicion of the government and immigration departments was a common misinformation theme: “some of us see [COVID] as a tactic for the government to access our documentation status and deport us”^34^. One Mexican male (age 45) in one US study^37^ noted: “When someone uploads something to Facebook then no-one believes in it 100%”; a Mexican female (age 33) was also quoted as saying “[I get my information] well through the news, TV, Facebook and all of that…not everything I see is credible”.

In a UK qualitative study^24^, participants who initially disregarded conspiratorial beliefs found it challenging to maintain their confidence that the rumours were untrue due to a number of factors: (i) receiving many social media messages about them; (ii) receiving messages about them from trusted others; (iii) feeing anxious; and (iv) being under lockdown conditions at home. Participants expressed confusion about which story to trust, and ongoing difficulty identifying information as misinformation and dismissing it. Similarly, another study^35^ reported that 79% of female Black Americans interviewed stated that they were confused by the COVID-19 information they’d accessed from any source: “Sometimes I feel unsure about the information that I’m receiving because it’s a lot of different things about it. Everybody’s not saying the same thing. So, I’m kind of unsure about what to believe”.^35^

### Social media impact on preventative health measures and vaccine intent

A small number of studies linked social media use with lower participation in preventative measures among migrants and ethnic minority groups. A UK/US survey study found vaccine hesitancy to be associated with informational reliance on social media and membership of an ethnic minority group^26^. A UK qualitative study reported that ethnic minority groups were influenced by anti-vaccine misinformation, including from social media.^24^ A UK qualitative study of precarious migrants found that among 23 participants who were hesitant about receiving a vaccine some participants described fears around theories based on misinformation, often originating from social media or word of mouth, with many describing feeling conflicted about which information sources to trust^29^. Community leaders from African, Caribbean, Asian and Eastern Mediterranean migrant groups in London, UK reported substantial COVID-19 vaccine hesitancy due to misinformation circulating on social media and word of mouth combined with a lack of accurate, translated and clear guidance^27^. Similarly, in a US qualitative study of Latino adults, some participants reported encountering a lack of knowledge accompanied by misinformation on social media causing them to dismiss preventative measures^34^. Another US study among Latino people reported that social media acted as a potential deterrent for following some public health measures to prevent infection by allowing people to observe the negative, guideline-breaking behaviours of others in social media posts^37^.

On the other hand, a large (8,001 participants) US/UK randomised controlled experiment^30^ found no significant differences in the response of different ethnic groups to misinformation in relation to vaccine intent. A large US/UK study^31^ found membership of an ethnic minority group was associated with reduced vaccine intention, a relationship which was significant in three out of four studies (p<0.001, n=3890; p=0.017, n=1663; p<0.001, n=2237). The relationship persisted even when use of legacy (print and broadcast media) and frequency of use of social media was controlled for. High levels of social media use was not associated with vaccine intent in any of the three studies exploring this relationship; however, high information reliance on social media was significantly associated with negative vaccine intent (p=0.028, n=2237), suggesting a reliance on social media for information can make users vulnerable to misinformation. This study did not include interaction terms between ethnicity and information reliance on social media, which could have indicated whether the effect of information reliance on social media on vaccine intent differs by ethnicity.

### Good practice in promoting information and countering misinformation

Evidence suggests the important role of strong connections with the local community to identify and counter misinformation and rumours by trusted and valued sources of information. Most studies recommended improving the accessibility of public health information for migrant and ethnic minority communities.^25 27 32 34 35 37-40 44^ For example, providing public health information in the media channel preferred by that group^34^, in multiple languages^24^, and using local, trusted voices delivering specific and targeted messages to counter fake news^24 34^. A strong interest in online, personalised information was identified^36 37^. Where social media was used to share personalised and culturally tailored public health information, it has a positive influence with good health knowledge, health seeking behaviours and vaccine intent^24-27 32 34-36 38-41 44^. Studies indicated the need for culturally tailored health messaging to ensure equitable health knowledge for improving vaccine intent and health seeking behaviours^27 34-36^.

More personalised means of health information communication was highlighted as a demand for informational reliance. A national US organisation which provides online health information tailored to the US Latino community found a high level of interest in their COVID-19 curated content, suggesting a strong demand for tailored and culturally relevant material^36^. In a new approach, ‘virtual patient navigators’, helpers working online, typically using messages to provide individually tailored health information, were made available to Latino migrants through a New York-based communication platform^32^.

Working through trusted sources was also emphasised. Providing accurate and tailored information about COVID-19 via trusted community members and organizations was suggested in a study of Black women aged 19-31 years in the US^35^.The study recommended that health professionals take an active role collaborating with the community to address inequities that Black women are experiencing in the pandemic^35^. Participants in a randomised controlled study to explore the impact of misinformation on vaccine intent on different populations groups reported finding videos on social media very engaging, especially when delivered in multiple languages by someone in a trusted profession (e.g., doctor/teacher/nurse)^30^.

Successful countering myths was reported in a UK study wherein the local council rapidly responded to fake news circulating in the local population (e.g., a rumour about children who test positive in school for COVID-19 being removed from the school and/or their parents until they test clear)^24^. Videos to refute the myth were swiftly posted online in both Urdu and Punjabi, and these were reported to be effective by members of the local population^24^. Additional studies report successfully countering misinformation using a network of patient navigators^32^ and community household surveys^24^. Social media use to communicate with family was also reported to be effective in challenging COVID-19 denial misinformation rumours through reporting of lived experience of COVID-19^34^.

## Discussion

Among migrant and ethnic minority populations in the UK, US, China, Jordan, Qatar, and Turkey we found evidence of consistent use of social media for COVID-19 information, including via WeChat, Facebook, WhatsApp, Instagram, Twitter, YouTube, which may stem from a difficulty in accessing COVID-19 information in their native languages or from sources they trusted. There were both positive and negative associations with social media use reported, with some evidence of circulating misinformation and social media use associated with lower participation in preventative health measures, especially vaccination intent, and finding that will be undoubtedly generalisable to multiple population groups. This is a rapidly evolving field of research, and data are limited, but our work highlights the considerable importance of social media platforms as a source of information and misinformation about COVID-19 for some migrant and ethnic minority populations during the pandemic. Whilst we know social media is used by many people, and misinformation has been circulating widely in the general population, it may be the case that those excluded from national public health responses and/or who faced specific barriers to accurate public health information and support may have been disproportionately impacted. Urgent actions and further research are now needed to better understand use of social media platforms for health information in different population groups, find effective approaches to tackling misinformation, and to seize on opportunities to make better use of social media platforms to support public health communication and improve vaccine uptake globally. Furthermore, the findings highlight the crucial role of locally trusted sources in identifying and tackling misinformation, and underscores the benefits of disseminating personalised and culturally relevant health messages, including via social media.

This review is the first attempt to synthesise global studies exploring the use and impact of social media on migrant and ethnic minority populations during the COVID-19 pandemic. However, it is limited by the availability and quality of the datasets available. We acknowledge the limited geographical scope of included studies, with 16 of 21 studies focused on migrant and ethnic minority populations residing in the UK and US and no data at all from low-income countries. We acknowledge that definitions and terms pertaining to migrants and ethnic minorities and social media are used inconsistently in research; this is an ongoing challenge within the field, which has previously been evidenced in similar reviews, and may mean we have missed papers. This was mitigated against by searching the published and grey literature more widely. In addition, we acknowledge that migrants and ethnic minorities are a highly diverse group with a range of health and socioeconomic situations making it hard to generalise; however there is evidence in several contexts that these populations may have been disproportionately impacted by the COVID-19 pandemic^13 15 45^.

The findings of our review have been confirmed by more recent studies. For example, a survey of migrants in Greece found their main source of information about the vaccine was via social media platforms and the internet in general, and that vaccine hesitancy was linked to a lack of adequate information and driven by fear, anxiety, exposure to negative news and misinformation^46 47^. In Turkey, a 2021 survey and feedback mechanism in refugee communities found information gaps, misconceptions, and rumours about COVID-19 vaccines circulating mainly by word of mouth and on social media, undermining health information^48^. In a recent study of Venezuelan migrants in Latin America, 70% said they had access to a mobile phone, with the main communication channels being WhatsApp and Facebook, yet half said they felt uninformed^49^. We also found that some migrants and ethnic minorities used diaspora media as a source of COVID-19 related information during the pandemic, which merits further consideration in terms of understanding how to better engage these groups in preventative health care and vaccination, and has been previously reported in studies as influencing views and beliefs around vaccination^50^. Misinformation on social media correlated negatively with vaccine intention and our findings align with other research in this area and will undoubtedly be relevant to many other population groups^2 7 29^. A recent study among migrants and nationals in Qatar acknowledged ‘personal research’ via social media as important to them for seeking information about COVID-19 vaccines, underlining the key role social media has in influencing people’s attitudes towards vaccine uptake^51^.

The European Centre for Disease Prevention and Control (ECDC) and other public health bodies have raised concerns around barriers to public health information among migrant populations and ethnic minority groups residing in Europe and other high-income countries during the pandemic^13 14^. Public health guidance in some countries was not initially tailored to the needs of migrant and ethnic minority groups^17 52-54^. A review of the availability of government produced risk communications across Council of Europe member states in June 2021 found only 48% (23/47) of countries translated COVID-19 information into at least one migrant language, with information on testing or healthcare entitlements in common migrant languages only found in 6% (3/47), suggesting individuals not able to access information in the host country language may have been excluded to some extent from governments’ public health messaging^17^. In Denmark, a series of qualitative interviews with migrants found that they felt uncertain regarding government guidance for COVID-19; although written material was translated into 19 languages, it was not effectively disseminated^55^. In Montreal, Canada, there were delays to publishing official multilingual fact sheets on COVID-19 guidelines, and information phone lines only operate in French and English; those who had arrived most recently, had lower language (French/English) ability or lower literacy had more difficulty accessing local COVID-19 information^56^. Lack of English or French language at the time of immigration to Canada were associated with lower rates of testing and higher percent positivity for COVID-19 in recently arrived adult immigrants and refugees^57^. A study among refugees and migrants in deprived areas in Greece found that migrants may have difficulties understanding public health messaging due to cultural and language barriers.^17 53 58^ Merely translating public health information is not likely to be sufficient; information needs to be tailored and targeted so it is conveyed in ways that resonate with the target population. A range of key resources and guidelines on risk communication and engagement strategies for COVID-19 public health responses, including vaccination, among marginalised populations globally are available, as well as a social media toolkit for healthcare practitioners (https://www.who.int/publications/m/item/a-social-media-toolkit-for-healthcare-practitioners---desktop)^59-61^. However, it will be vitally important that the lessons learned around communication of public health information to marginalised groups during the pandemic are meaningfully carried forward.

Where social media is used to share personalised and culturally tailored public health information, it has a positive correlation with good health knowledge, health seeking behaviours and vaccine intent^36 41^ Our research shows the need for culturally tailored health messaging to ensure equitable health knowledge and to improve vaccine uptake, by accurate public health messaging through trusted sources of information^27 34-36^. We make a number of recommendations for policy and practice (Table 4), which include the need for systematic monitoring of information and attitudes circulating on social media^62^, as well as timely rebuttal of misinformation from trusted professionals. Several resources are now available to support addressing misinformation about COVID-19 vaccines as well as fostering demand for vaccines.^63-65^

**Table 4:**
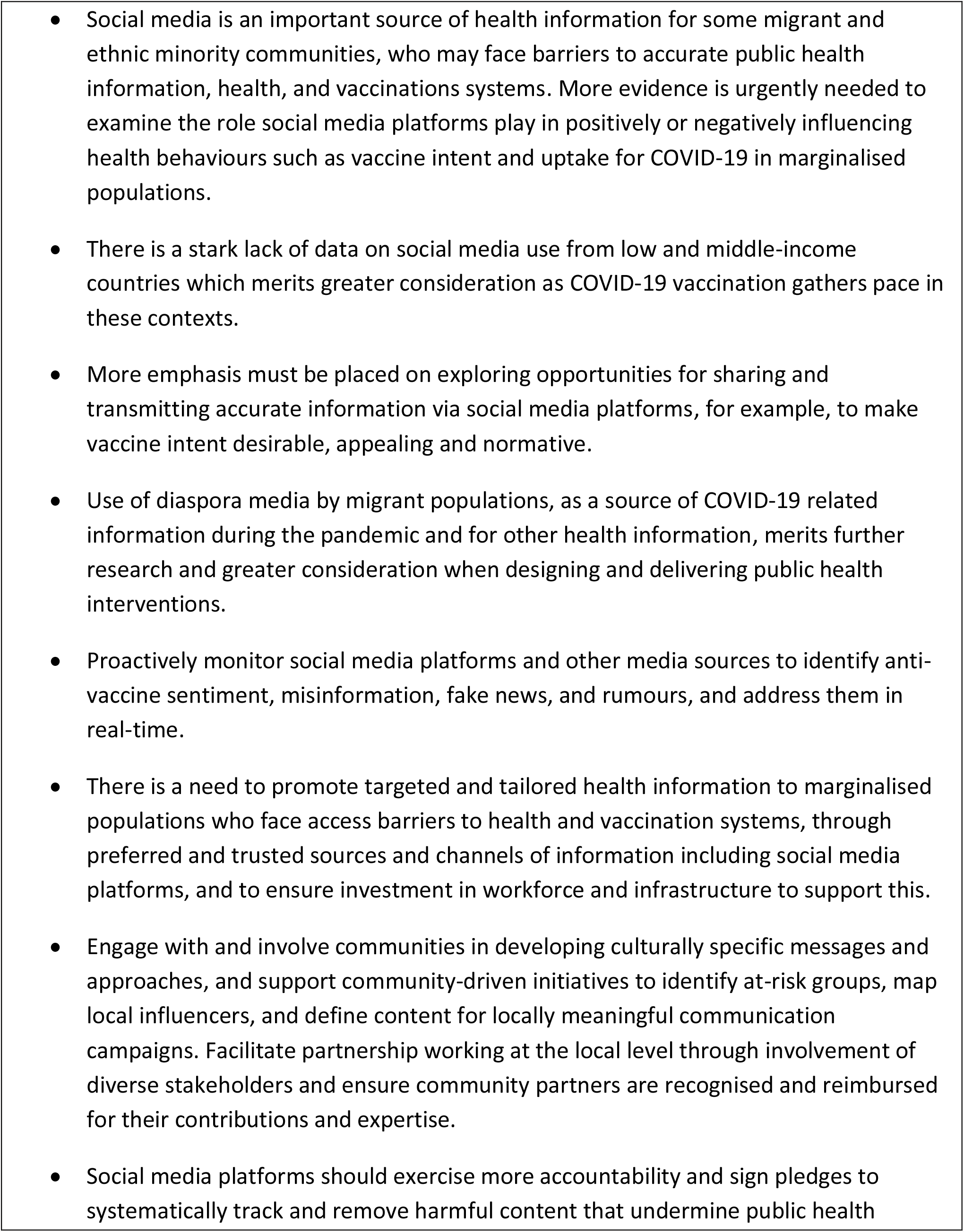
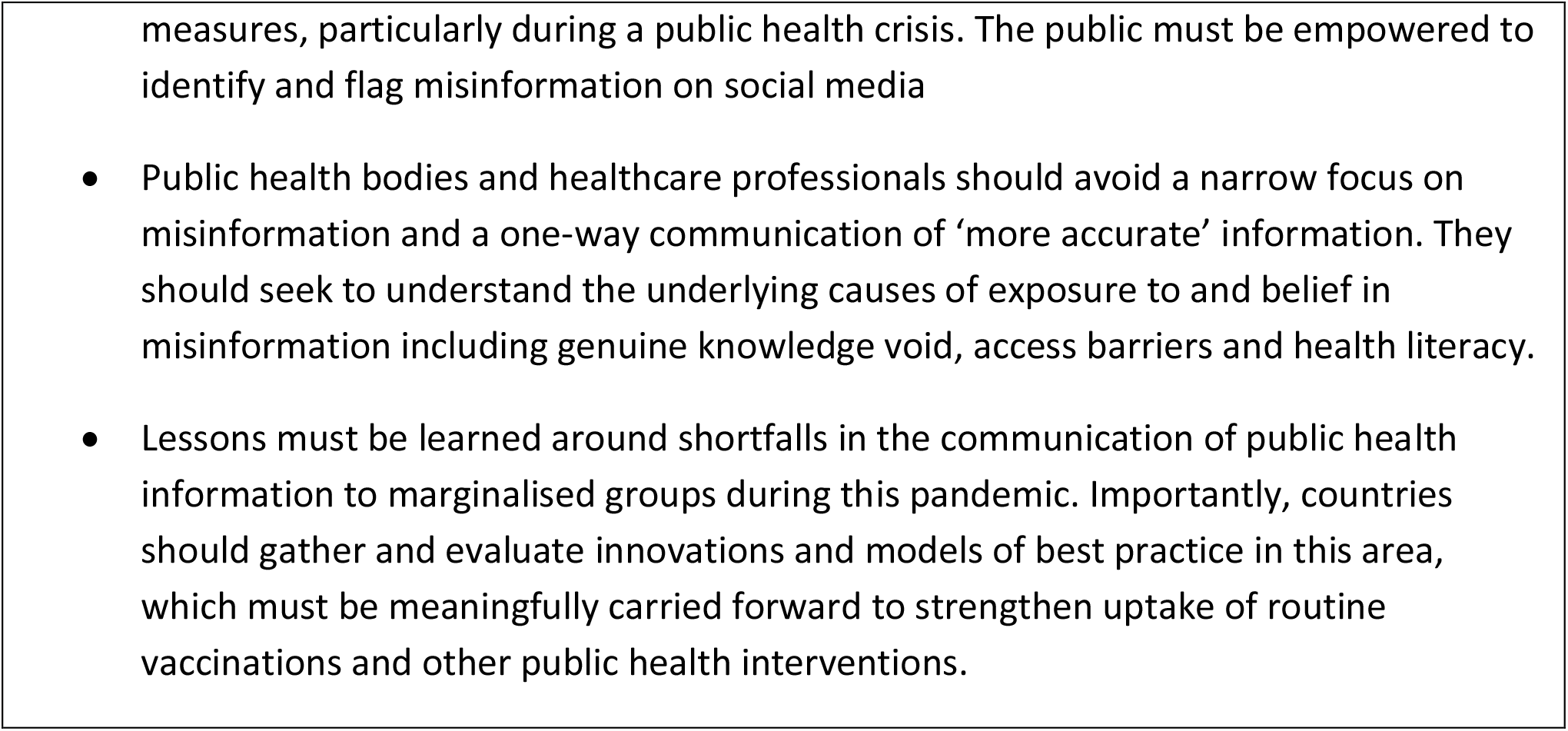
Key messages and recommendations.

There is a stark lack of data on social media use from low and middle-income countries, which merits greater consideration as COVID-19 vaccination gathers pace in these contexts. In addition, more evidence is needed to examine the role social media platforms play in positively or negatively influencing health behaviours such as vaccine intent and uptake for COVID-19 in all populations (including other excluded groups eg, homeless, internally displaced people/IDPs). Social media is an important source of health information for some migrant and ethnic minority communities and tacking misinformation needs to be done using this medium given the lack of trust in government messaging in some of these communities^66^. Our findings are consistent with those of others working in this field, which show that social media can have a crucial role in disseminating health information, tackling infodemics and misinformation^4^. There is an opportunity now to more effectively use social media to make vaccine intent desirable, appealing and normative among migrants and ethnic minority groups. There is an urgent need to address infodemic-related challenges in a rapidly changing information environment, including real-time monitoring of social media messages and misinformation and the development of online tools to fight disinformation, with a focus on collecting stratified population data to enable targeted and tailored responses. Robust interventions relying on behavioural science to tackle misinformation using social media and evaluations are a plausible next step to address immunisation challenges for COVID-19 vaccines but also routine vaccines. Building trust in public health messaging, identifying information gaps, finding innovate ways of disseminating health information, and detecting and responding to misinformation as it emerges remain a priority for public health^66 67^.

## Data Availability

All data used in the systematic review are appropriately referenced and available online in the sources cited.

## Ethics statements

Not applicable.

## Patient consent for publication

Not applicable.

## Funding and acknowledgements

This study was funded by the National Institute for Health Research (NIHR300072) and Academy of Medical Sciences (SBF005\1111). LPG, SH, and AFC are funded by the NIHR (NIHR300072); AFC and SH are funded by the Academy of Medical Sciences (SBF005\1111). SH acknowledges funding from the Novo Nordisk Foundation (Mobility– Global Medicine and Health Research) and the World Health Organization. AD and SEH are funded by the Medical Research Council (MRC/N013638/1). KR is funded by the Rosetrees Trust (M775). MR is funded by an NIHR In-Practice Clinical Fellowship (NIHR 302007). JC is funded by an NIHR In-Practice Clinical Fellowship (NIHR300290). The views expressed are those of the author(s) and not necessarily those of the NHS, the NIHR, or the Department of Health and Social Care. The funder of the study had no role in study design, data collection, data analysis, data interpretation, or writing of the report.

## Contributors

The study was conceptualised by SH, and the protocol and research question were developed by SH, LPG and MRP. Searches were developed by MRP and LG, with input from SH and SEH. Screening was done by LPG and MRP. Data extraction and analysis was done by LPG and MRP, with input from SH. The first draft of the manuscript was produced by LG, MRP, and SH, and developed with KH and TV, who all contributed to interpretation of the results. All authors commented upon and approved the final manuscript. SH is guarantor of this study.

## Disclaimer

The views expressed are those of the author(s) and not necessarily those of the NHS, the NIHR, or the Department of Health and Social Care. The funder of the study had no role in study design, data collection, data analysis, data interpretation, or writing of the report.

## Supplementary materials

**Online supplement 1:** Boolean search terms.

|  |  |
| --- | --- |
| Migrant and ethnic minorities | Ancest* OR Diaspor* OR ethnic* OR Ethnoc* OR Ethnog* OR "Identity politics" OR Ingroups OR outgroups OR Intersectionality OR Kinship OR "Minority group*"~3 OR "minority population*"~2 OR minorities OR Multicultu* OR Polyethnic* OR "Population genetics" OR Race OR races OR racial OR Tribe* OR latino*) OR AB:(Ancest* OR Diaspor* OR ethnic* OR Ethnoc* OR Ethnog* OR "Identity politics" OR Ingroups OR Outgroups OR Intersectionality OR Kinship OR "Minority group*"~3 OR "minority population*"~2 OR minorities OR Multicultu* OR Polyethnic* OR "Population genetics" OR Race OR races OR racial OR Tribe* OR latino*) OR "afro american*"~3 OR BAME OR latino* OR roma OR romani OR refugee* OR immigrant* OR "migrant" OR "displaced person" OR "displaced persons" OR "social determinant*"~2 OR "latin population" OR "latin group*" OR "people of color" OR "people of colour" |
| Social media | social media OR social network OR online communit* OR online discuss* OR online communicat* OR online post OR messag* OR chat OR media OR |
|  | misinformat* OR disinformat* OR malinformat* OR fake new* OR twitter OR<br>whatsapp OR tweet OR post OR instagram OR reddit OR weibo OR sina OR<br>youtube OR tiktok OR snapchat OR pinterest OR likee OR sharechat OR<br>discord OR kuaishou OR wechat OR weixin OR qq OR telegram OR quora OR<br>mobile app OR blog OR podcast OR hashtag OR antivax* OR vaccine<br>hesitanc* OR web 2.0 OR online |

## References

1. WHO. Health topics/Infodemic. Available from: https://www.who.int/health-topics/infodemic#tab=tab_1.

2. N. Puri, et al. Social media and vaccine hesitancy: new updates for the era of COVID-19 and globalized infectious diseases. Human Vaccines & Immunotherapeutics, 16 (11) (2020), pp. 2586–2593.

3. H. Oi-Yee Li, et al. YouTube as a source of information on COVID-19: a pandemic of misinformation? BMJ Global Health, 5 (2020).

4. Tsae S-F, et al. What social media told us in the time of COVID-19: as scoping reviews. Lancet Digit Health 2021; 3:e175–94..

5. C.H. Basch, et al. What do popular YouTube videos say about vaccines? Child Care Health Dev, 43 (4) (2017), pp. 499–503.

6. Larson, Heidi J. “The biggest pandemic risk? Viral misinformation.” Nature, vol. 562, no. 7726, Oct. 2018, p. 309. Gale Academic OneFile, link.gale.com/apps/doc/A573035610/AONE?u=anon∼d201a883&sid=googleScholar&xid=01fdad7a..

7. Wilson SL, Wiysonge C. Social media and vaccine hesitancy. BMJ Global Health 2020; 5 (10) https://gh.bmj.com/content/5/10/e004206.

8. Roozenbeek J, Schneider CR, Dryhurst S, et al. Susceptibility to misinformation about COVID-19 around the world. Royal Society Open Science 2020;7(10):201199. doi: 10.1098/rsos.201199.

9. American Press Institute. Race, Ethnicity, and the Use of Social Media for News. How Millennials Use Technology to Get News 2015. https://www.americanpressinstitute.org/publications/reports/survey-research/race-ethnicity-social-media-news/.

10. Gorman D, Bielecki K, Willocks L, Pollock K. A qualitative study of vaccination behaviour amongst female Polish migrants in Edinburgh, Scotland. Vaccine. 2019;37(20):2741–7.

11. World Health Organization. 10 global health issues to track in 2021. Spotlight. https://www.who.int/news-room/spotlight/10-global-health-issues-to-track-in-2021.

12. Cinelli M, Quattrociocchi W, Galeazzi A, et al. The COVID-19 social media infodemic. Scientific Reports 2020;10(1):16598. doi: 10.1038/s41598-020-73510-5

13. Hayward SE, Deal A, Cheng C, et al. Clinical outcomes and risk factors for COVID-19 among migrant populations in high-income countries: A systematic review. Journal of Migration and Health 2021;3:100041. doi: https://doi.org/10.1016/j.jmh.2021.100041

14. ECDC. Reducing COVID-19 transmission and strengthening vaccine uptake among migrant populations in the EU/EEA. Technical Report. ECDC: Stockholm, 2021..

15. Hargreaves S, Hayward S, Noori T, McKee M, Kumar B. COVID-19: Counting migrants in. Lancet 2021; 398..

16. Turkish Red Crescent Society CCs, Fatma Nur Bakkalbaşı, Onurcan Ceyhan, Tenadi Gölemerz. COVID-19 Rumour Tracking Report: Turkish Red Crescent Society (TRCS) in Collaboration with International Federation of Red Cross and Red Crescent Societies (IFRC), 2021.

17. Maldonado BMN, et al. Engaging the vulnerable: a rapid review of pubic health communication aimed at migrants during the COVID-19 pandemic in Europe J Migr Health 2020; 1:100004. https://pubmed.ncbi.nlm.nih.gov/33447830/.

18. Rowland-Pomp M, Hargreaves S, Goldsmith LP, et al. The impact of social media and misinformation on migrant and ethnic minorities populations and their response to the COVID-19 pandemic: a systematic review. PROSPERO 2021 CRD42021259190, 2021.

19. Page MJ, McKenzie JE, Bossuyt PM, et al. The PRISMA 2020 statement: an updated guideline for reporting systematic reviews. BMJ 2021;372:n71. doi: 10.1136/bmj.n71

20. World Health Organization. COVID-19 Global literature on coronavirus disease database https://www.who.int/emergencies/diseases/novel-coronavirus-2019/global-research-on-novel-coronavirus-2019-ncov.

21. Ouzzani M, Hammady H, Fedorowicz Z, et al. Rayyan—a web and mobile app for systematic reviews. Systematic reviews 2016;5(1):1–10. doi: 10.1186/s13643-016-0384-4

22. Protogerou C, Hagger MS. A checklist to assess the quality of survey studies in psychology. Methods in Psychology 2020;3 doi: 10.1016/j.metip.2020.100031

23. Critical Appraisal Skills Programme. CASP Qualitative Checklist. 2018 doi: https://casp-uk.net/casp-tools-checklists/

24. Lockyer I, Rahman, Dickerson, Pickett, Sheldon, Wright, McEachan, Sheard. Understanding Covid-19 misinformation and vaccine hesitancy in context: Findings from a qualitative study involving citizens in Bradford, UK, 2020.

25. Paul E, Steptoe A, Fancourt D. Attitudes towards vaccines and intention to vaccinate against COVID-19: Implications for public health communications. The Lancet Regional Health Europe 2021;1:100012–12. doi: 10.1016/j.lanepe.2020.100012

26. Allington(a) D, McAndrew S, Moxham-Hall V, et al. Coronavirus conspiracy suspicions, general vaccine attitudes, trust and coronavirus information source as predictors of vaccine hesitancy among UK residents during the COVID-19 pandemic. Psychol Med 2021:1–12. doi: 10.1017/S0033291721001434

27. Crawshaw AF, Deal A, Rustage K, et al. What must be done to tackle vaccine hesitancy and barriers to COVID-19 vaccination in migrants? J Travel Med 2021;28(4):taab048. doi: 10.1093/jtm/taab048

28. Vekemans T. Crisis and Continuation: The Digital Relocation of Jain Socio-Religious Praxis during the COVID-19 Pandemic. Religions 2021;12(5):342–42. doi: 10.3390/rel12050342

29. Deal A et al. Strategies and action points to ensure equitable uptake of COVID-19 vaccinations: a national qualitative interview study to explore the views of undocumented migrants, asylum seekers, and refugees. J Migr Health 2021; 4: 100050..

30. Loomba S, de Figueiredo A, Piatek SJ, et al. Measuring the impact of COVID-19 vaccine misinformation on vaccination intent in the UK and USA. Nature Human Behaviour 2021;5(3):337–48. doi: 10.1038/s41562-021-01056-1

31. Allington(b) D, McAndrew S, Moxham-Hall VL, et al. Media usage predicts intention to be vaccinated against SARS-CoV-2 in the US and the UK. Vaccine 2021;39(18):2595–603. doi: 10.1016/j.vaccine.2021.02.054

32. Behbahani S, Smith CA, Carvalho M, et al. Vulnerable Immigrant Populations in the New York Metropolitan Area and COVID-19: Lessons Learned in the Epicenter of the Crisis. Acad Med 2020;95(12):1827–30. doi: 10.1097/ACM.0000000000003518

33. Campos-Castillo C, Laestadius LI. Racial and Ethnic Digital Divides in Posting COVID-19 Content on Social Media Among US Adults: Secondary Survey Analysis. J Med Internet Res 2020;22(7):e20472–e72. doi: 10.2196/20472

34. Cervantes L, Martin M, Frank MG, et al. Experiences of Latinx Individuals Hospitalized for COVID-19: A Qualitative Study. JAMA Netw Open 2021;4(3):e210684–e84. doi: 10.1001/jamanetworkopen.2021.0684

35. Chandler R, Guillaume D, Parker AG, et al. The impact of COVID-19 among Black women: evaluating perspectives and sources of information. Ethn Health 2020:1–14. doi: 10.1080/13557858.2020.1841120

36. Despres C, Aguilar R, McAlister A, et al. Communication for Awareness and Action on Inequitable Impacts of COVID-19 on Latinos. Health Promot Pract 2020;21(6):859–61. doi: 10.1177/1524839920950278

37. Moyce S, Velazquez M, Claudio D, et al. Exploring a rural Latino community’s perception of the COVID-19 pandemic. Ethn Health 2020:1–13. doi: 10.1080/13557858.2020.1838456

38. Viswanath K, Bekalu M, Dhawan D, et al. Individual and social determinants of COVID-19 vaccine uptake. BMC Public Health 2021;21(1):818–18. doi: 10.1186/s12889-021-10862-1

39. Woko C, Siegel L, Hornik R. An Investigation of Low COVID-19 Vaccination Intentions among Black Americans: The Role of Behavioral Beliefs and Trust in COVID-19 Information Sources. J Health Commun 2020;25(10):819–26. doi: 10.1080/10810730.2020.1864521

40. Wang C, Tian Q, Zhao P, et al. Disease knowledge and attitudes during the COVID-19 epidemic among international migrants in China: a national cross-sectional study. Int J Biol Sci 2020;16(15):2895–905. doi: 10.7150/ijbs.47075

41. Hamadneh S, Hamadneh J, Amarin Z, et al. Knowledge and attitudes regarding Covid-19 among syrian refugee women in Jordan. Int J Clin Pract 2021;75(5):e14021–e21. doi: 10.1111/ijcp.14021

42. Alabdulla M, Reagu SM, Al-Khal A, et al. COVID-19 vaccine hesitancy and attitudes in Qatar: A national cross-sectional survey of a migrant-majority population. Influenza Other Respir Viruses 2021;15(3):361–70. doi: 10.1111/irv.12847

43. Danish Refugee Council. COVID-19 Impact on Refugees in South East Turkey. 2020 doi: https://data2.unhcr.org/en/needs-assessment/1432

44. (R4V) RI-aCP. Information and communication needs assessment - U-Report Uniendo Voces Regional Poll, 2021.

45. Buikema AR, et al. Racial and ethnic disparity in clinical outcomes among patients with confirmed COVID-19 infection in a large US electronic health record database. Lancet EClinMed 2021; Sept 101075..

46. Hellenic Red Cross. CEA. Survey on the migrant’s population informatio needs, regarding health issues (COVID-19) https://communityengagementhub.org/resource/survey-on-the-migrants-population-information-needs-regarding-health-issues-covid-19/.

47. Hellenic Red Cross. Perceptions survey on Covid-19 vaccination, Greece. https://communityengagementhub.org/resource/perceptions-survey-on-covid-19-vaccination-greece/.

48. COVID-19 rumour tracking report. Ankara: Turkish Red Crescent Society; 2021 (https://communityengagementhub.org/wp-content/uploads/sites/2/2021/10/Rumour-tracking-report-2021.pdf, accessed 11 January 2022).

49. Only half of refugees and migrants from Venezuela feel informed, survey finds. Panama City: Coordination Platform for Refugees and Migrants from Venezuela; 2020 (https://www.r4v.info/sites/default/files/2021-06/CWC%20EN.pdf,, accessed 11 January 2022).

50. Ganczak M, Bielecki K, Drozd-Dabrowska M, Topczewska K, Biesiada D, Molas-Biesiada A et al. Vaccination concerns, beliefs and practices among Ukrainian migrants in Poland: a qualitative study. BMC Public Health. 2021;21 (1):93. doi: 10.1186/s12889-020-10105-9.

51. Alabdulla M, Reagu SM, Al-Khal A, Elzain M, Jones RM. COVID-19 vaccine hesitancy and attitudes in Qatar: A national cross-sectional survey of a migrant-majority population. Influenza Other Respir Viruses. 2021;15(3):361–70.

52. Patel P, Hiam L, Orcutt M, Burns R, Devakumar D, Aldridge R, et al. Policy brief: Including migrants and refugees in the British government’s response to COVID-19. 2020.

53. Doctors of the World. An Unsafe Distance: The Impact of the COVID-19 Pandemic on Excluded People in England. 2020.

54. Kondilis E, Papamichail D, Mc Vann, Carruthers E, Veizis A, Orcutt M, Hargreaves S. The impact of the COVID-19 pandemic on refugees and asylum seekers in Greece: a retrospective analysis of national surveillance data, 2020. Lancet EClinMed 2021; 37: 100958: https://doi.org/10.1016/j.eclinm.2021.100958.

55. Institut for Menneskerettigheder. Corona rammer skævt - etnicitet og smitte. Copenhagen, Denmark; 2020.

56. Cleveland, J, Hanley, J, Jaimes, A, Wolofsky, T., 2020. Impacts de la crise de la COVID-19 sur les «communautés culturelles »montréalaises: Enquête sur les facteurs socioculturels et structurels affectant les groupes vulnérables. Montréal: Institut universitaire SHERPA.

57. Guttmann, A, Gandhi, S, Wanigaratne, S, Lu, H, Ferreira-Legere, L, Paul, J, et al., 2020. COVID-19 in Immigrants, Refugees and Other Newcomers in Ontario: Characteristics of Those Tested and Those Confirmed Positive, as of June 13, 2020. ICES, Toronto, ON.

58. Cholera, R, Falusi, OO, Linton, JM., 2020. Sheltering in place in a xenophobic climate: COVID-19 and children in immigrant families. Pediatrics 146 (1).

59. Conducting community engagement for COVID-19 vaccines: interim guidance, 31 January 2021. Geneva: World Health Organization; 2021 (https://apps.who.int/iris/handle/10665/339451, accessed 11 January 2022).

60. COVID-19 immunization in refugees and migrants: principles and key considerations: interim guidance, 31 August 2021. Geneva: World Health Organization; 2021 (https://www.who.int/publications/i/item/covid-19-immunization-in-refugees-and-migrants-principles-and-key-considerations-interim-guidance-31-august-2021.

61. Good practice guidance for risk communication and community engagement (RCCE) for refugees, internally displaced persons (IDPs), migrants and host communities particularly vulnerable to COVID-19 pandemic. London: British Red Cross; 2020 (https://communityengagementhub.org/wp-content/uploads/sites/2/2020/06/Practical-Guidance-RCCE-Refugees-IDPs-Migrants.pdf.

62. The Vaccine Confidence Project. Media Monitoring Report UK COVID-19 01 June to 31 July 2021. London: LSHTM https://www.vaccineconfidence.org/research-feed/social-media-conversations-and-attitudes-in-the-uk-towards-covid-19.

63. Vaccine misinformation management field guide: guidance for addressing a global infodemic and fostering demand for immunization. New York: United Nations Children’s Fund; 2020 (https://vaccinemisinformation.guide).

64. Understanding the infodemic and misinformation in the fight against COVID-19: digital transformation toolkit. Washington (DC). Pan American Health Organization; 2020 (https://iris.paho.org/bitstream/handle/10665.2/52052/Factsheet-infodemic_eng.pdf?sequence=16.

65. How to address COVID-19 vaccine misinformation. Atlanta (GA): United States Centers for Disease Control and Prevention; 2021 (https://www.cdc.gov/vaccines/covid-19/health-departments/addressing-vaccine-misinformation.html.

66. Vandrevala, Hendy, Hanson, Alidu & Ala (2022). Unpacking COVID-19 and Conspiracy Theories in the UK Black Community. Pre-print..

67. Vandrevala T, et al. “It’s possibly made us feel a little more alienated’: how people from ethnic minority communities conceptualise COVID-19 and its influence on engagement with testing. Jan 3, 2022 https://doi.org/10.1177%2F13558196211054961.

68. Statista. https://www.statista.com 2021 [cited 2021 17th Sept 2021]. Available from: https://www.statista.com/topics/1164/social-networks/#dossierKeyfigures accessed 17th Sept 2021 2021.

